# Risk analysis of chewing betel quid among diabetic patients from the northeastern part of Bangladesh

**DOI:** 10.1101/2020.01.15.20017731

**Authors:** Zafrul Hasan, Md. Rafiul Islam, Md. Soyib Hasan, Miah Mohammad Sakib, Md. Sifatul Islam, Md. Belal Chowdhury, Saifuddin Sarker, Md. Rakibul Islam, Mohammad Abul Hasnat, Lalith Mohon Nath, Md. Waseque Mia

**Affiliations:** Dept. of Biochemistry and Molecular Biology, Shahjalal University of Science and Technology, Sylhet-3114, Bangladesh; Sylhet Diabetic Hospital, Sylhet, Bangladesh; Dept. of Biochemistry and Molecular Biology, University of Dhaka, Dhaka-1000

**Keywords:** Betel quid, Areca nut, Hyperglycemia, Diabetes mellitus, RBS

## Abstract

**Background:** Betel quid (BQ) chewing is a common habit and a means of social interaction among the northeastern peoples of Bangladesh. Though this habit integrating in their daily life without knowing its toxic effect. Areca nut, which is one of the main components of BQ and may responsible for this addiction. Here, we assess to see how BQ chewing habit influence hyperglycemia among diabetic patients with respect to their lifestyle.

**Methodology:** Random blood sugar (RBS) test was evaluated from a total of 961 diabetic patients. Behavioral data associated with their daily lifestyle were collected from August 2018 to February 2019 from Sylhet Diabetic Hospital, Bangladesh. Student’s t-test, ANOVA and Fisher’s exact test were used to assess the RBS status between BQ chewer and non-chewer patients.

**Results:** Higher RBS was found in BQ chewer patients than non-chewer (mean ± SEM, 263.3 ± 4.768 vs. 251.0 ± 5.915mg/dl). Interestingly, it is significantly higher in raw areca nut user than dry nut (mean ± SEM, 278.0 ± 8.790 vs. 252.1 ± 6.835 mg/dl) only from BQ chewer group, suggesting that the habit of chewing raw nut may contribute to more hyperglycemic effect among diabetic patients. BQ habit enhances higher RBS level among smoker, non-smoker and patient’s having walking habit. In addition, BQ habit significantly influence to have high RBS in patients with family history with diabetes. Lack of awareness being diabetes have also been observed significantly in BQ chewer patients, while a higher level of RBS was seen in BQ group, who work in different sectors with sitting activities.

**Conclusions:** Diabetic patients who chew betel quid are more prone to keep higher hyperglycemic. Utmost attention should be taken to discourage the use of BQ for proper management of diabetes control.

## Introduction

Chewing areca nut (major component) with others ingredients (e.g. betel leaf, slaked lime, and may contain tobacco) generally known as betel quid (BQ), is considered the fourth most common source of addiction, after caffeine, nicotine and alcohol [1,2]. The addiction of BQ among chewers is due to the presence of areca nut [3]. Indeed, BQ chewing is increasing alarmingly worldwide as a mean of social interaction, predominantly in Asian countries but also seen in North America, Africa, Australia, and Europe, because of immigrants [1,4]. The contributing factors of its widespread use are due to alertness, less fatigability and ecstatic feeling among the users because areca nut (locally known as supari or betel nut) contains various psychoactive alkaloids, mainly arecoline which bind to GABA receptors of brain and trigger the effects [3,5]. It is becoming evident that this particular bioactive ingredient interferes with insulin mediated glucose uptake by cells and associated with type II diabetes along with other diseases in betel nut users (including coronary artery and metabolic disease) [2,6]. In addition, the most common accompaniment for chewing areca nut is the leaf of *piper betel* (locally known as paan or betel leaf) in BQ, which is known for its antidiabetic potential that has been, demonstrated *in vivo* experiments [7,8]. Therefore, it is of interest how areca nut in the presence of betel leaf in BQ influences hyperglycemic state among diabetic patients in population level is still enigmatic.

Chewing BQ is one of the popular traditional habits in Bangladesh among men and women equally [4]. However, this habit is more prevalent in Sylhet division (the northeastern part of Bangladesh), may be due to the long history of cultural influence of nearby Indian boarder area. This population has also another tendency of chewing different form of nut, raw and/or dry in their BQ preparation. Interestingly, frequency of diabetic patients in Sylhet, aged 35 years or more are high, like other division (south, southeastern and central part, where peoples are wealthier and educated with hypertension) of Bangladesh [9]. Therefore, in this population-based study, we aim to assess any association of chewing BQ and life style (e.g. smoking, physical activities, age, awareness and others) on hyperglycemic status among diabetic patients, which have never been explored in this region of Bangladesh.

## Methods

### Study population

All the participants for this population-based study were from northeastern part of Bangladesh, Sylhet. People of Sylhet region, of various age groups are traditionally habituated with betel leaf and areca nut chewing than other region of Bangladesh and diabetes prevalence also high among this local people [9]. General people who are less educated with relaxed lifestyle and source of income mostly of foreign remittance as one of the members from each household live in abroad. This place is geographically and environmentally very suitable region (subtropical climate and rich terrain) of growing *areca catechu* tree for areca nut and *piper betel* plant for betel leaf.

### Study design and random blood sugar test

A total of n=961 patient’s blood samples and their behavioral data from August 2018 to February 2019, (RBS mg/dl, median 237; IQR, 170-321.5) were collected, whom visited Sylhet Diabetic Hospital from morning to noon for random blood sugar test. RBS testing is a powerful tool for people with diabetes to assess how well the disease is being managed and this test can be used to measure the blood glucose level regardless to the time of meal. We did not account any reference value above or below the threshold level of RBS rather all range of sugar level were collected as the patients have already diagnosed with diabetes and visiting hospital for routine check. Duplication was excluded using unique id provided by hospital as well patients name. Patients who chew betel leaf with areca nut as dry and/or raw form considered as betel quid positive chewer, otherwise betel quid negative chewer. Data was excluded from the analysis of patients having only betel leaf or areca nut chewing habit along. Blood was drawn from each participant and allowed for clotting at room temperature followed by centrifuged at 3000 rpm for 10 minutes. Separated serum was used for RBS test according to manufacturer’s instructions (Human Diagnostic, Germany) with a semi-automated biochemistry analyzer (Humalyzer 3000, USA).

### Questionnaire data

All the participants of diabetic patients were given a consent form for their approval and after getting their informed consent, a standard questionnaire set was circulated among the study participants to answer. Information on sociodemographic data were collected by trained medical stuff, which contained responder’s name, id, age, sex, physical activity, body weight, smoking, alcohol intake, type of diabetes, family history, occupation, age of first time diagnosis, betel leaf, betel nut (raw or dry form) and cellular number using one-to-one interview. This population-based study was received ethical approval by the ethic committee of school of life sciences, Shahjalal University of Science and Technology (SUST), Bangladesh.

### Statistical analysis

All Statistical analyses were performed using Prism 6 software (GraphPad). A *p* value <0.05 indicated by asterisk (significant), otherwise *p* value > 0.05 indicated by ns (not significant). Unpaired t-test or student’s t-test (two-tailed) was done to assess the differences between independent variables. To determine any differences among the groups One-way ANOVA was performed. To examine the significance of association between the two kinds of variables, Fisher’s exact was performed. All through values were represented as mean ± standard error of the mean (SEM), otherwise noted.

## Results

### Chewing BQ and random blood glucose among diabetes patients

To see the effect of chewing BQ on random blood glucose (RBS, mg/dl) level among diabetic patients, we enrolled, n=961 individuals with their consent. RBS generally assess how well the disease is being managed and the test can be used to measure glucose level at any time to identify hyperglycemia. Though BQ chewer group was shown higher RBS level, Fig 1 (mean ± SEM, 263.3 ± 4.768) than non-chewer (mean ± SEM, 251.0 ± 5.915) but not reached statistical significance. Previous study have shown areca nut which is one of the main components of BQ is responsible for hyperglycemia [2]. Interestingly the local people of Sylhet region are very familiar with adding raw nut in their BQ preparation with other ingredients as it has mild head spinning and mouth aroma effect rather than dry nut. Almost half of the studied population use raw nut in their quid and the proportion of raw vs. dry nut chewer was 45.6% and 54.4% respectively. It is not practical to chew only betel leaf or nut alone, rather local folks prefer to use in combined form and thus the issue guide us to see the effect of chewing raw and dry nut on RBS level. Patients who chewed raw nut in BQ had significantly higher RBS (mean ± SEM, 278.0 ± 8.790 vs. 252.1 ± 6.835 mg/dl) than those with dry nut user (Fig 2). In addition, patients who chewed both raw and dry nut in BQ (mean ± SEM, 261.6 ± 9.726), no significant difference of RBS was found either with raw or dry nut chewer group of patients (Fig 2). Taken together, this result demonstrating that raw nut may play an important role in keep rising blood sugar more in diabetic patients and this subtle outcome cannot be seen in dry and both nut user. This is may be due to the presence of psychoactive alkaloids in raw nut, which may be lost or compromised its spectrum during the drying or other processes before placing in BQ preparation.

**Fig 1.**
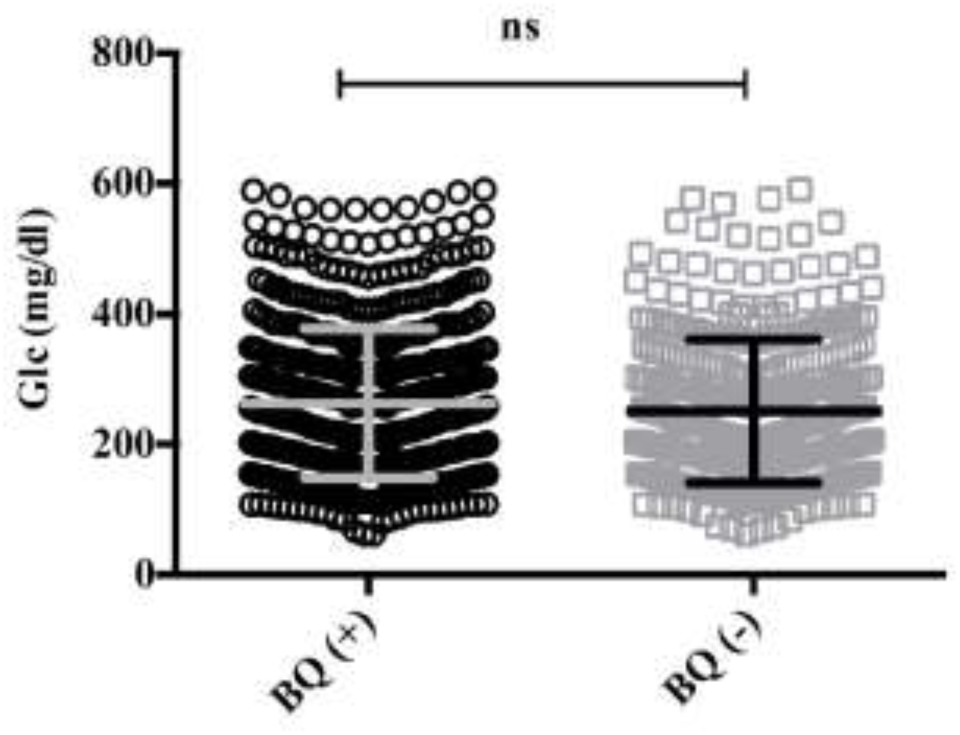
BQ chewing effect on diabetes patients. Patients with BQ chewer habit shown higher RBS level (mean ± SEM, 263.3 ± 4.768) than non-chewer (mean ± SEM, 251.0 ± 5.915), though it did not reach statistical significant. ns, not significant using student-t test where *p* value > 0.05 is significant.

**Fig 2.**
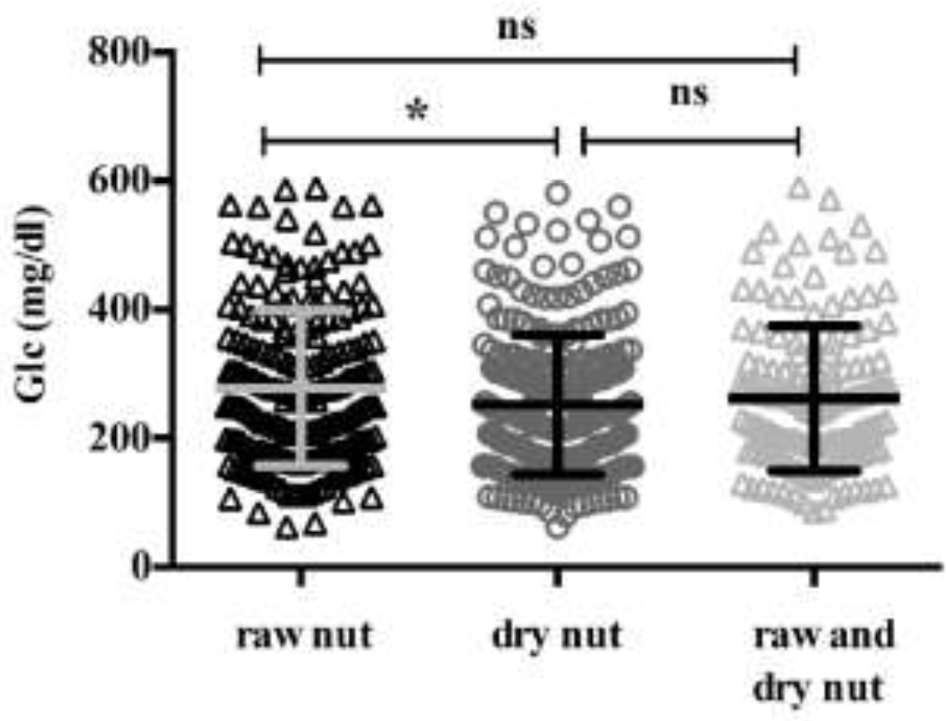
Effect of raw and dry form of areca nut in BQ preparation among diabetes patients. Higher RBS (mean ± S.E.M, 278.0 ± 8.790 mg/dl) was found in raw nut chewer than that of dry nut chewer patients (mean ± S.E.M, 252.1 ± 6.835 mg/dl), which is statistically significant (*p* < 0.0186), using student-t test.

### Effect of smoking on BQ chewer of diabetic patients

Smoking habit is a well-known and independent risk factor in many diseases, including diabetes [10,11]. To see any association between BQ and smoking habit among the diabetic patients, we split the data between BQ chewer and non-chewer with their smoking habit. Surprisingly, no additive effect of smoking on BQ chewer was found, while significantly higher RBS was found in nonsmoker BQ chewer group (S1 Fig). From our initial analysis of Fig 1 and 2, RBS was significantly higher in BQ chewer having raw nut, which prompted us to look the effect of smoking on BQ chewers, with practice of adding raw and/or dry nut in their quid preparation. Interestingly, BQ chewers with raw nut group have shown significantly higher RBS than non-BQ chewers (mean ± SEM, 291.5 ± 19.89 vs 232.4 ± 15.48) within the smoker patients (Fig 3a). It is also important to note that same trend of significant higher RBS was found in BQ chewers with raw nut group even in nonsmoker patients (Fig 3b). Thus, this result indicating that, presence of raw nut in BQ preparation may be responsible for keeping higher RBS level in diabetic patients, regardless their smoking habit.

**Fig 3.**
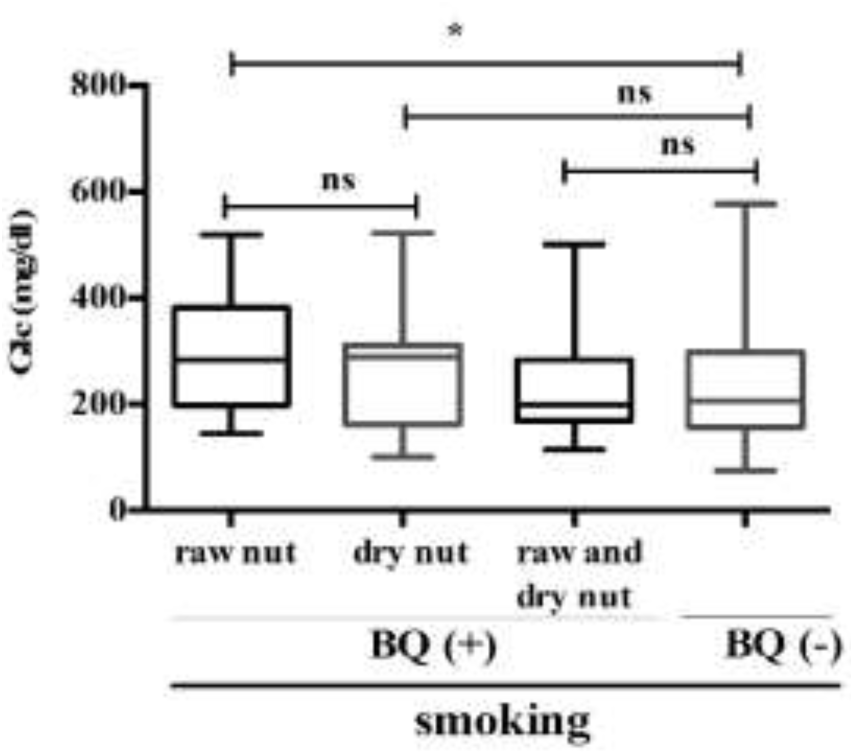
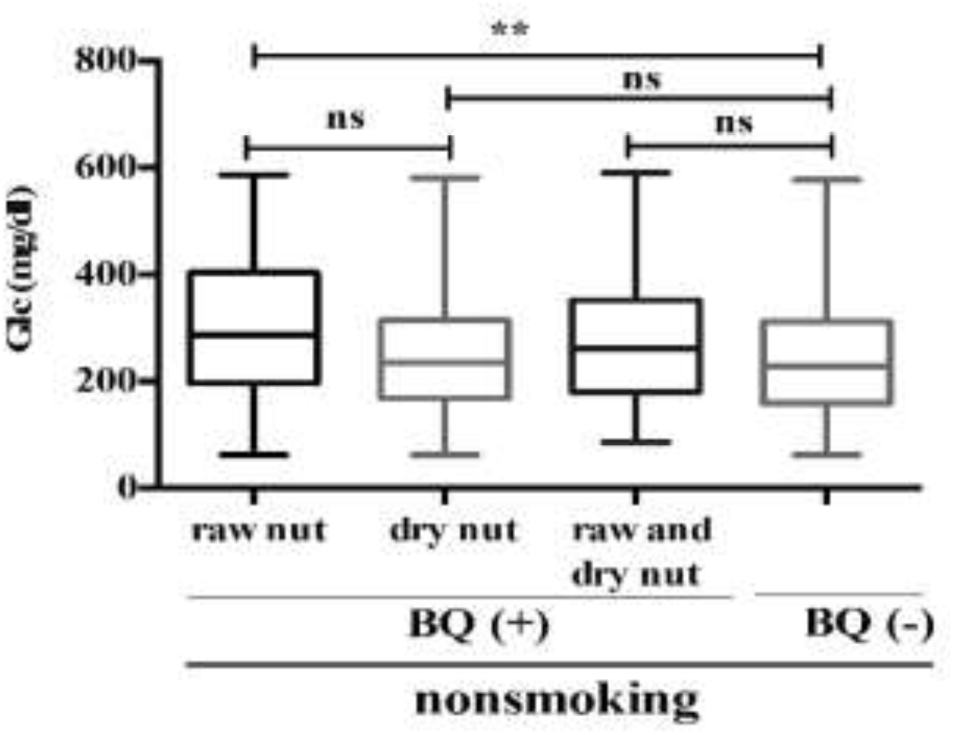
Effect of BQ chewing on diabetes patients having smoking habit. **(a)** Patients with smoking habit, significantly higher RBS was found in raw nut BQ chewer (mean ± SEM, 305.0 ± 17.19) with respect to no BQ chewer group (mean ± SEM, 245.8 ± 9.718). **(b)** Patients with nonsmoking habit, significantly higher RBS was found in raw nut BQ chewer (mean ± SEM, 291.5 ± 19.89) with respect to no BQ chewer group (mean ± SEM, 232.4 ± 15.48). *p* value <0.05 indicated by asterisk (significant), otherwise *p* values are > 0.05 indicated by ns (not significant), using student-t test.

### Physical activities on BQ chewer of diabetes patients

It is reported that physical activities has lowering effect on blood glucose level [12,13], which guide us to see how BQ chewing habit influences on physical activity toward patients RBS. Even though it seems that BQ has no impact on patients with or without physical activity by fisher’s exact test (Fig 4), interestingly RBS level was higher (mean, 297 mg/dl) in BQ group even with physical activity. This is an indication that BQ may have exacerbated effect toward rising RBS level, even people who has walking habit.

**Fig 4.**
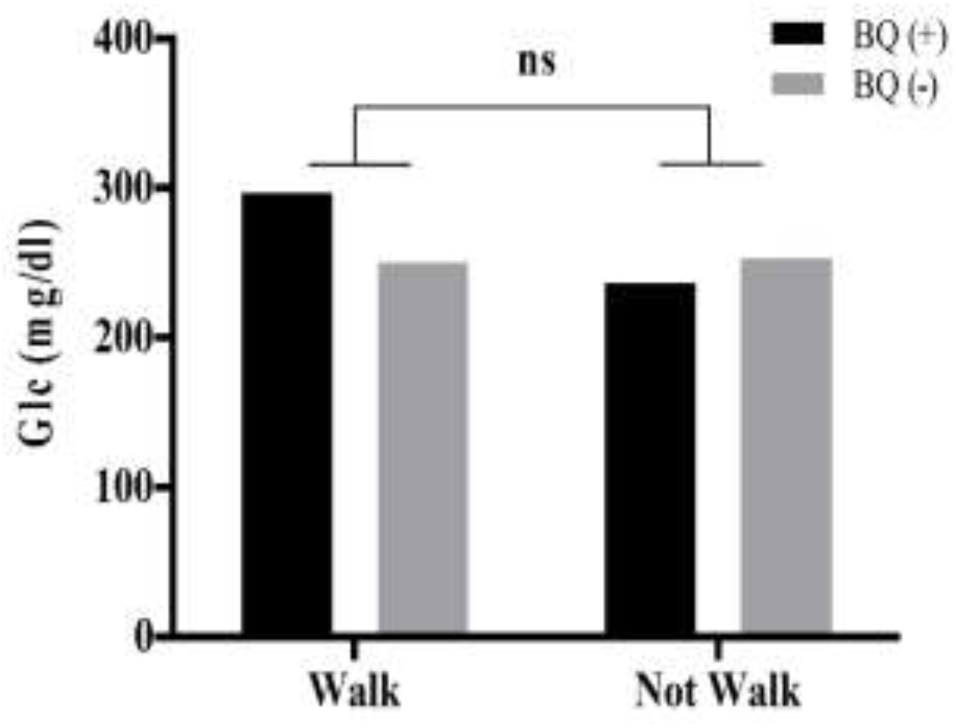
Impact of BQ chewing on diabetes patients having walking habit. BQ chewer diabetes patients with walking habit has no impact on lowering RBS level, with respect to not walking BQ chewer patients, which is statistically not significant by Fisher’s exact test.

### Family history and BQ chewing habit of diabetes patients

Family history is one of the major risk factors to developed diabetes at any stage of individual’s life [14-16]. Current understanding of heritability or genetic predisposition information is not enough but also need to address the possible risk factors that are associated with our daily life, such as BQ habit. In this study, we have defined family history, at least one of the family members have diabetes, which may be either, father / mother / sibling or any two or all of them. Thus, our results indicating that patients with family history of diabetes have more probability of being higher RBS with BQ chewer habit than non-chewer (mean ± SEM, 258.3 ± 7.432 vs. 238.3 ± 6.414), which is significant (Fig 5). Interestingly, without any family history of diabetes, this BQ habit has also tiny effect on RBS level (Fig. 5), supporting that BQ has negative impact on diabetic patients.

**Fig 5.**
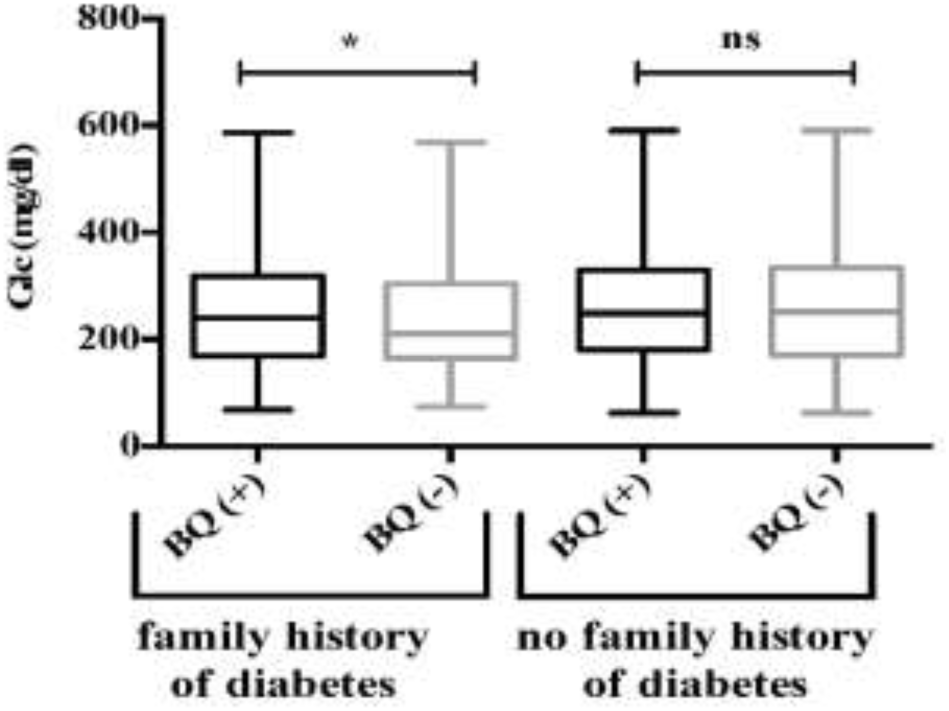
Influence of BQ chewing habit on diabetes patients with family history. Higher RBS was found in diabetes patients having BQ habit who has family history of diabetes, which is significant using student-t test (*p*< 0.05).

### Lack of awareness of being diabetes among BQ chewers

BQ users have been experienced reinforcing effects including, reduce tension, euphoria, sense of well-being, and increased capacity to work, due to the psychoactive properties of its major components, areca nut. [17,18]. Considering the previous literatures and our result from this studied diabetic population have shown that BQ habit may put the individual in euphoric and bit confident that they may not have problem with blood glucose, which, however, delayed diagnosis of having diabetes (Fig. 6). It could be possible the individuals suffering with hyperglycemia little early before the diagnosis which is statistically significant between chewer and non-chewer group (*p* < 0.0006). It is conceivable that this lack of awareness may be due to presence of areca nut or supari, because people believe that it has medicinal potential with long history of use, as herbal drug [19].

**Fig 6.**
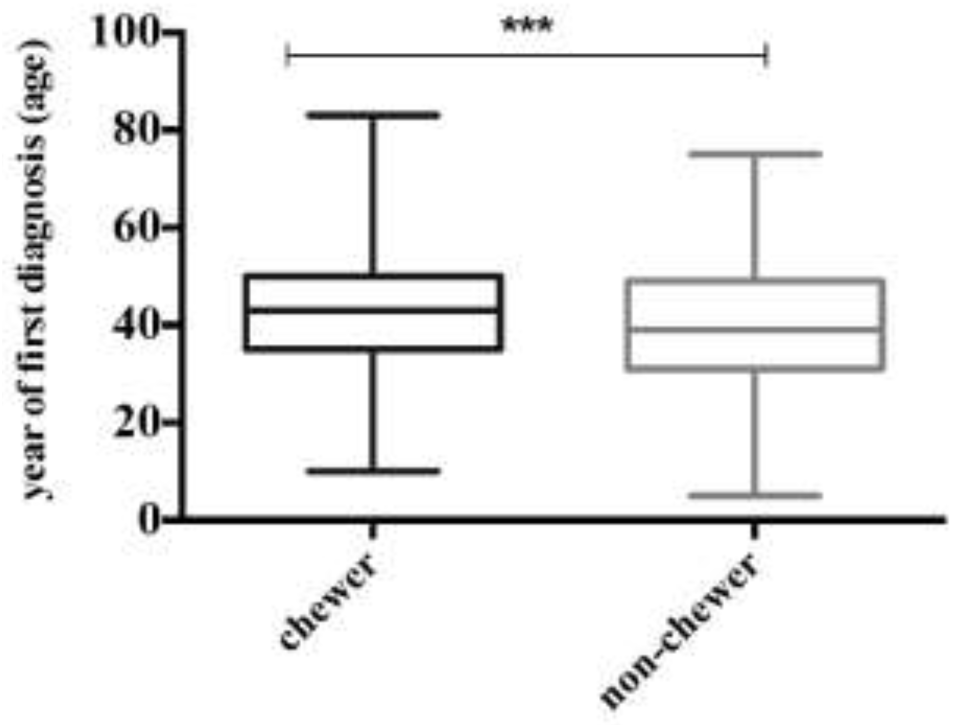
Awareness of being hyperglycemic between BQ chewer vs. non-chewer. BQ chewer peoples have tendency of diagnosis with diabetes at the late age with respect to non-chewer, which is statistically significant using student-t test (*p* < 0.0006).

### Influence of BQ chewing on various occupational people

We observed people with various occupations have BQ chewing habit irrespective of their education and income status in Sylhet. Several studies indicate that diabetes are more prevalent in those peoples who has occupation with sedentary activity [20,21]. Here we explore the effect of BQ on diabetes patients having various occupational backgrounds (e.g. agriculture, business, driver, office worker, teacher, housewife and others). Like other reported study, we have also seen the significantly higher level of RBS (Table 2) in BQ chewer group who work in different offices with sitting job (mean ± SEM, 283.6 ± 16.63 vs. 227.6 ± 11.52 mg/dl). Noteworthy, higher RBS was also seen in other occupations with BQ chewer group than non-chewer except the housewife, because this group might be always occupied themselves with many family issues throughout the day, which therefore, promoting physical activities.

**Table 1.** Glucose level as RBS among various aged group diabetes patients with and without BQ habit.

| Age group (years) | Prevalence rate (%) | | Glucose (mg/dl)<br>mean ± SEM | | *p* (<0.05) |
| --- | --- | --- | --- | --- | --- |
|  | chewer | non-chewer | chewer | non-chewer |  |
| <30 | 28.8 | 71.2 | 229.0 ± 34.46 | 258.8 ± 21.03 | *ns* |
| 30-39 | 52.2 | 47.8 | 271.0 ± 13.10 | 248.6 ± 14.24 | *ns* |
| 40-49 | 64.5 | 35.5 | 253.1 ± 8.68 | 234.9 ± 11.45 | *ns* |
| 50-59 | 69.4 | 30.6 | 261.4 ± 7.844 | 252.7 ± 11.75 | *ns* |
| 60-69 | 62.5 | 37.5 | 278.7 ± 13.60 | 267.4 ± 15.21 | *ns* |
| >70 | 68.6 | 31.4 | 273.0 ± 17.32 | 247.3 ± 18.30 | *ns* |

BQ = betel quid, *ns* = not significant

**Table 2.** RBS level among various occupants with diabetes having BQ chewing habit or not.

| Occupation | Glucose (mg/dl)<br>mean ± SEM | | *p* (<0.05) |
| --- | --- | --- | --- |
|  | chewer | non-chewer |  |
| Agriculture | 274.1 ± 19.69 | 252.4 ± 31.52 | *ns* |
| Business | 266.3 ± 14.23 | 250.7 ± 15.62 | *ns* |
| Driver | 287.1 ± 39.07 | 244.4 ± 47.02 | *ns* |
| Housewife | 256.3 ± 5.900 | 260.7 ± 8.772 | *ns* |
| Office worker | 284.2 ± 13.91 | 227.6 ± 11.52 | ** |
| Teacher | 260.0 ± 20.13 | 244.2 ± 19.64 | *ns* |
| Others | 272.2 ± 53.14 | 252.1 ± 37.01 | *ns* |

Data are expressed as chewer vs. non-chewer with significant *p* values (<0.05) indicated by
asterisks, otherwise *p* values are > 0.05 by student-t test. BQ = betel quid, Others = unemployed,
students, and no information, *ns* = not significant.

## Discussion

In this population-based study on northeastern part of Bangladesh, Sylhet, we attempt to assess how the BQ chewing practice influence hyperglycemic level on diabetic patients considering various confounder factors associated with daily life styles (e.g. physical activity, smoking, family history, age, occupation and awareness). One of our keen interests of focus was, using of areca nut in form of raw and dry in their BQ preparation. Local people of this region of various ages prefer to have BQ with raw and/or dry nut all through their working and leisure time. Our preliminary analysis indicates that the prevalence rate of chewing BQ tendency becoming higher in people with age and interestingly RBS level also being high in these older age group people with diabetes Table 1. We believe this may be due to the presence of areca nut in BQ, because it contains psychoactive addictive substances (e.g. arecoline and other alkaloids) [2,5] and also cannot rule out the other factors, such as family milieu and cultural influences towards getting this BQ habit as a mean of socialization with others. This is alarming because the main workforce people (30-50 years) are affected by this BQ habit without knowing its long term toxic effect on them, since the toxicity of BQ chewing is usually mild and unobserved [18].

Although chewing habit of areca nut in the form of betel quid (BQ) is very old and cultural practice, which is intensely integrated in people’s daily life, not only in Asia but also in different immigrants dominated parts of the world [1,4,22]. Previous study has reported that chewing areca nut is associated with diabetes [2] and we have seen in our study that diabetic patients who chew raw nut in BQ preparation has significantly higher RBS than that of dry nut chewer (Fig 2). Interestingly this difference could not be seen between BQ chewer vs non-chewer (Fig 1) irrespective of used form of nut, indicating that it is imperative to evaluate the ingredients and it’s form in BQ preparation separately. Indeed, we also did not find any significant difference of RBS in smoker patients with or without BQ chewing habit, but could see significant difference in nonsmoker patients S1 Fig. Again after considering the used form of nut in BQ in smoker and nonsmoker, significant difference was appeared between chewer with raw nut vs. non-chewer diabetes patients but not with dry nut chewer (Figs 3a and 3b), implying that raw form of nut user are in more prone to elevated RBS, without knowing its silent hyperglycemic effect.

A healthy lifestyle such as physical activity or walking can keep one’s blood glucose level in target range, while lack of such activity or sedentary lifestyles are highly associated with diabetes [23]. Analysis of our dataset, observed that BQ chewing has adverse impact on keeping higher blood glucose level even the patients with walking activity (Fig 4). It should be noted that we could not get the exact information of patients walking nature, i.e. how long, frequency, what time of the day, indoor (treadmills), outdoor and others. Some people have very healthy lifestyle, while being diagnosed with diabetes, because of predisposition with family history of diabetes or genetic association. Our analysis also revealed that patients with family history has significantly higher RBS with BQ chewing habit (Fig 5), and collectively these results are suggesting that the BQ habit may contribute to keep the diabetes patient in hyperglycemic state subtly and which, however, been overlooked by medical community, because of lack of proper literature.

We have seen people of various ages of Sylhet region have BQ chewing habit Table 1 and believe that it is good for health, particularly for digestion. Arecoline, a major psychoactive alkaloid of areca nut in BQ preparation, trigger mild euphoric effect and sense of well-being [3,5]. Remarkably our data indicate that BQ chewer are significantly less aware of being hyperglycemic and perform diabetes test later than of non-chewer people (Fig 6). In addition, people with different occupations have also been seen with this BQ chewing habit i.e. agriculture, businessman, driver, office worker, housewife, and, teacher, may be due to the euphoric effect by areca nut. Indeed, our result with good agreement with other study expressed that significantly higher RBS was identified people with sedentary occupation, like office work, who used to sit in their chair with mouth of BQ Table 2.

This study is not without any limitations and one of the major confounder factors that we could not integrate whether the patients were under medication at the time of enrollment. Sometimes patients actually reluctant to answers lot of questions, specifically the nature of medication use, other than their doctor. Another few factors, like household income, level of education, food habit (junk or healthy), tobacco use, sample size, are the few possible confounders need to be addressed in future extended study including other areas of Bangladesh. One of the key strength of this study, that we included several factors associated with socioeconomic and lifestyles, which were lacking in some of the previous studies by other groups [2,23]. Moreover, this study population was quite homogeneous population as they were almost locally born in Sylhet, not like other big divisions where peoples coming from different part of Bangladesh for work purposes.

## Conclusions

Taken together, this population-based study suggesting that diabetes patients with chewing BQ habit are associated with elevated level of blood glucose. Indeed, attending medical doctor in the diabetes hospital has not aware that BQ could be one of the reasons of hyperglycemia because of lack of proper literature and knowledge. Therefore, BQ habit may have literally negative impact on diabetes management and treatment. Utmost attention should be given towards BQ as this habit becoming an addiction and integrating silently in our social life and discourage as strongly as other addiction like tobacco. Thus, medical community should convey this information in the society for proper management of BQ mediated hyperglycemia and should highlight the need for more research of this addictive disorder.

## Data Availability

Data will be available with few restrictions stipulated by Shahjalal University of Science and Technology (SUST), Sylhet, Bangladesh

## Acknowledgments

We thank Dr. A Z Mahbub Ahmed, Superintendent, Sylhet Diabetic Hospitals, Sylhet, Bangladesh for his approval and support of this study. We wish to thank the technicians, students of BMB, SUST for their generous help in sampling and data collection. Authors are thankful to all participants for blood sample and their participation in this study.

## Author Contributions

Conceived and designed the study: ZH, MRI, MSH, MMS, and MSI. Sampling and data collection: MRI, MSH, MMS, and MSI. Analyzed the data: ZH. Suggestion and technical help: MWM and MAH. Wrote the paper: ZH. Reviewed/edited the manuscript: ZH and MRI.

## Notes

### Competing Interest Statement

The authors have declared no competing interest.

### Funding Statement

No funding

